# Identifying High Priority Ethical Challenges for Precision Emergency Medicine - A Nominal Group Study

**DOI:** 10.1101/2023.02.23.23286334

**Authors:** Christian Rose, Emily Shearer, Isabela Woller, Ashley Foster, Nicholas Ashenburg, Ireh L. Kim, Jennifer Newberry

## Abstract

**OBJECTIVE:** Precision medicine is a rapidly progressing avenue to providing the right care to the right patient at the right time and spans all medical fields and specialties. However, given its reliance on computation and timely, accurate information, actualizing precision medicine within the emergency department and its “anyone, anywhere, anytime” approach presents unique challenges which could exacerbate disparities rather than improve care.

**METHODS:** We performed a qualitative, nominal group technique study of emergency physicians with prior knowledge of precision medicine concepts to identify high priority ethical concerns facing the implementation of precision medicine in the emergency department.

**RESULTS:** Twelve emergency physicians identified 91 ethical concerns which were organized into a framework with three major themes: values, privacy, and justice. The framework identified the need to address these themes across three time points of the precision medicine process: acquisition of data, actualization in the care setting, and after effects of its use.

**CONCLUSIONS:** Precision medicine may help to improve the quality of care provided in the emergency department, but significant hurdles exist. Our framework helps to identify high-yield ethical concerns that could serve as focus areas for future research and policy in order to guide the effective implementation of precision medicine in the emergency department.

## Introduction

Historically, medical treatments have been tailored to the average patient. However, few individuals look or feel like the average patient–each has their own risk factors, needs, and ideal outcomes. Precision medicine aims to address this problem by moving from a one-size-fits-all model toward one of providing the “right care to the right person at the right time” through the use of big data like genetics and wearable technologies.^1–5^ However, even the most advanced, targeted therapies are limited by biases that already exist in the US healthcare system.^6,7^ Differential care, even when “evidence based”, can result in worsened gaps between the haves and the have-nots, disproportionately burdening already marginalized communities.^8^ For emergency physicians, who serve in the safety net for American healthcare, precision medicine’s “right care, right patient, right time” must be balanced with our “anyone, anywhere, any time” mantra and the practical and ethical challenges that this setting presents.

To date, most precision medicine initiatives have focused on the utilization of genetics to better understand or treat diseases. Much has been made of determining the right medication for the right person (e.g., warfarin dosing for those with a CYP mutation) or when to start cancer screening based on genetic profiles (e.g., colorectal screening for CHD1).^9^ It has become clear, however, that genetics alone cannot explain much of the variation in outcomes across the population.^2,10^ The effect of behavioral factors like diet and exercise have been well-established, and studies have also begun to illuminate the effect of social and environmental determinants of health like access to healthcare resources or housing, which may be responsible for over half of health outcomes.^11,12^ As such, there has been a broadening approach to precision medicine that does not rely solely on genetics, but also home monitoring, social media and social networks, as well as patient-generated data like diet, daily steps, exercise, or heart rate variability.^13,14^

While useful when available, not all patients can afford these tools and services. Furthermore, “precise” treatments rely on the accuracy of this data, which must be built into data sets and shared. Few comprehensive data sets that incorporate all of these data exist, and even fewer have been carefully evaluated for missing or inaccurate data which could bias results.^15^ Current data sets suffer from a lack of representation from some populations while simultaneously over-representating chronically-ill patients or those presenting to research institutions.^16^ Data that were collected where patients are provided differential care might inadvertently support antiquated practices^17–21^ due to the association of outcomes with variables like the ability to pay for a higher level of care. Though precision approaches aim to limit quality gaps and provide more equitable care, decisions that are made without the full picture of their context might inadvertently exacerbate systemic biases.^7^

The emergency department setting offers additional unique hurdles for the growth of precision medicine. Little patient-generated data are readily available at the point of care. Highly important features like code status are haphazardly available among the sickest patients. Nevertheless, emergency care providers strive to provide better care, and to actualize the benefits of a truly learning healthcare system. How can this safety net, responsible for a growing and significant portion of acute care in America,^22,23^ begin to approach the problems that inherent to the implementation of precision medicine so that it might offer real-time, equitable care for patients with the widest spectrum of disease in medicine and a microcosm of the healthcare system?

Given the continued growth yet limited scope of precision medicine in the emergency department, our primary goal was to identify themes and gaps in current research by using a rigorous consensus technique to identify ethical questions and concerns of precision medicine relevant to emergency medicine practice. This manuscript presents the resulting consensus framework.

## Methods

### Study Design and Setting

We employed a modified nominal group technique (NGT), a consensus methodology widely used in emergency medicine, to identify high priority research questions relevant to the implementation of precision medicine in the emergency medicine setting and arrive at a consensus framework.^24–31^

NGT is a four-step process in which stakeholders generate and then prioritize research ideas (Figure 1). NGT was well-suited for our research question as our aim was to generate a comprehensive set of ethical questions and concerns which could then be organized and ordered. This is in contrast to other frequently used consensus techniques such as the Delphi technique, which are designed instead to reach a consensus opinion on one particular question.^24^ NGT has previously been used successfully to identify research priorities in the field of Emergency Medicine.^25,27,32–35^

**Figure 1:**
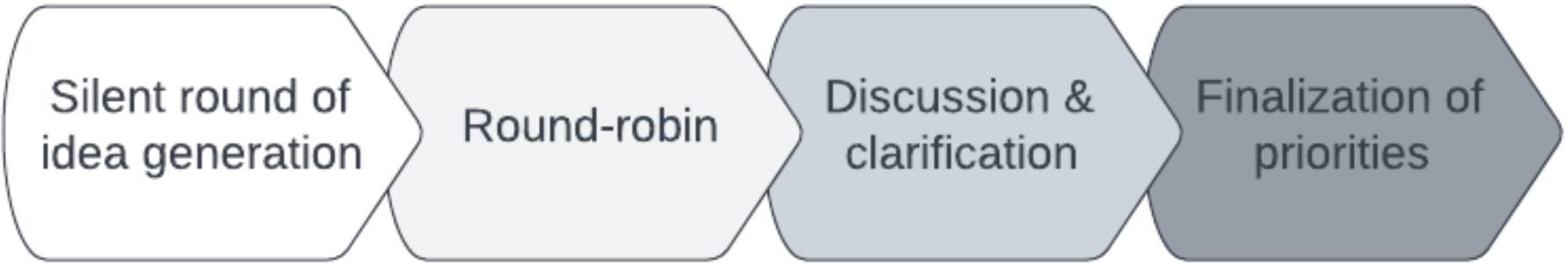
Flow chart of the NGT process.

### Selection and Participants

The ideal NGT session group size is greater than six - with the ideal between nine and twelve - participants,^32^ with the ideal between nine and twelve.^32^ Participants were recruited via email to the Society of Academic Medicine Ethics Committee listserv; the email invitation also asked recipients for referral and sharing to others who may be interested. A convenience sample of the first fourteen respondents to the email were registered as potential participants in the NGT group to meet the ideal group size and allow for scheduling conflicts. Ultimately, twelve emergency medicine physicians with working knowledge of precision medicine topics joined our NGT session. Participants represented a spectrum of sub-specialties (education, ethics, pediatrics, diversity, equity, and inclusion, and informatics) across various stages of training (resident, early-, mid- and late-career) and practice settings (county hospital, community hospital, academic center, and integrated managed care consortium) (Appendix 1) in order to provide a range of emergency medicine practice perspectives.

Participants took part in the NGT session during the American College of Emergency Physicians annual conference in Boston, October of 2021. The process was moderated by the lead researcher of the group (CR), and notes were taken by two separate researchers (ES and MG).

#### NGT Process

##### Phase 1: Silent idea generation

The first step of NGT is a round of silent idea generation. In this phase, after being given an initial prompt, participants silently write down what they believe to be high priority topics or concerns. It is imperative that this phase is silent, as this allows each member of the group to write their initial thoughts without influence from other members. Our prompt asked participants to identify what they thought were priority research topics for the implementation of precision medicine initiatives in the emergency medicine setting.

##### Phase 2: Round-robin

After individual idea generation, participants then shared their ideas with the group in a round-robin format. NGT has the advantage of not allowing any one particular voice to outweigh others, by asking each participant to share their complete list of ideas in-turn during the round robin phase, rather than ad-hoc discussion, thus minimizing potential bias.^36,37^

##### Phase 3: Discussion and clarification

After the round-robin format, participants undertook a 30-minute discussion and clarification of topics. In this phase, participants were encouraged to ask other members for clarification of topics generated as needed. Cross-cutting themes were identified, and duplicate topics consolidated. Key topics were then placed on moveable cards for prioritization and further thematic grouping.

##### Phase 4: Finalization of research priorities

Lastly, participants finalized the topics they believed represented priority areas for research. In traditional NGT, this is the ranking phase, where members are allocated votes to generate a ranked list of research priorities. Instead, participants categorized concerns by overarching themes and came to consensus on those distinctions.

## Results

During phases one and two of the NGT process, 82 questions were generated (Complete list in Appendix 2) with the average participant contributing 6.8 (+/- 2.9) questions. After refinement and separation of cross-cutting themes and consolidation of duplicate topics in phase three, a list of 68 final questions were retained.

In phase four, those questions were mapped into common thematic realms based on the nature of the ethical issue and where in the precision medicine process the issue was likely to occur (Tables 1-3). The result of this mapping was a 3 by 3 matrix consisting of ethical concerns grouped into the domains of values, privacy and justice as related to their temporal relationship to the precision medicine process, specifically in the acquisition of data, the actualization of precision medicine in practice, and the after-effects of the process (Figure 2). This matrix represents the product of our expert panel consensus.

**Table 1:**
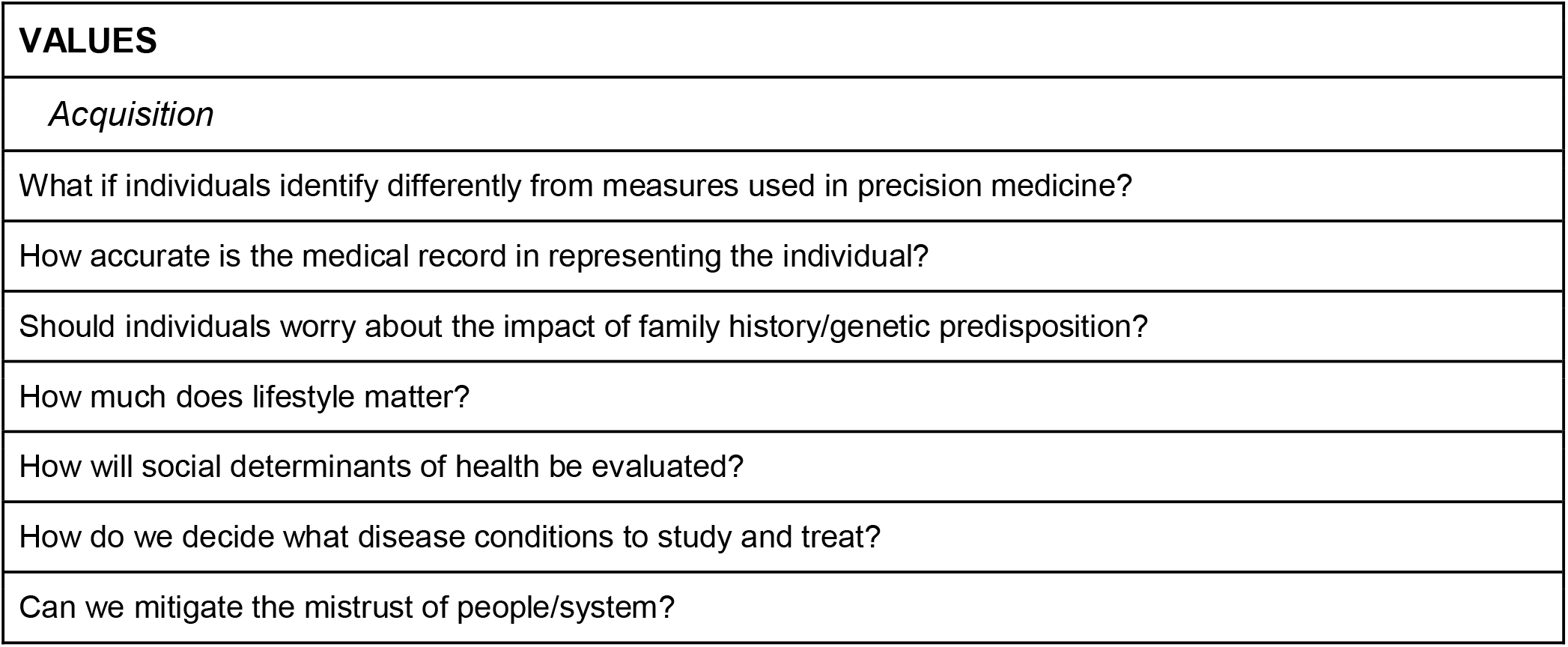
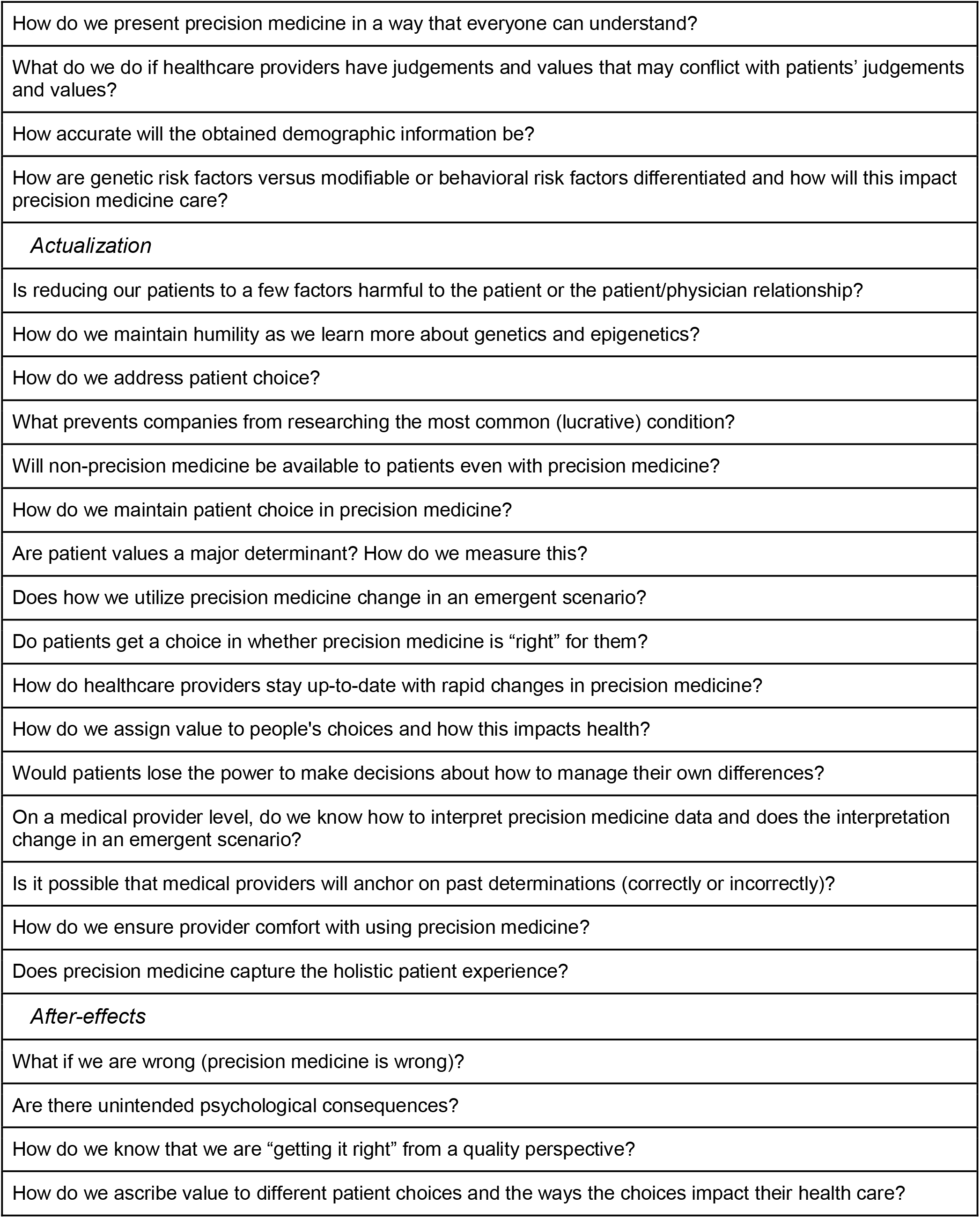
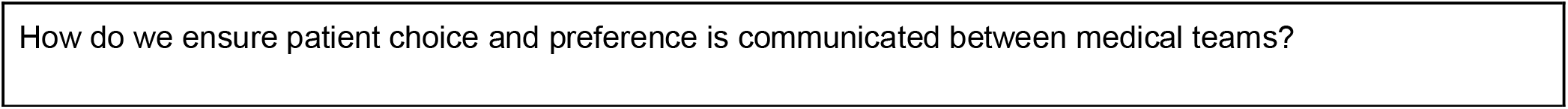
Participant concerns related to how the unique values of patients, providers, and the healthcare system will be incorporated into precision medicine.

**Table 2:**
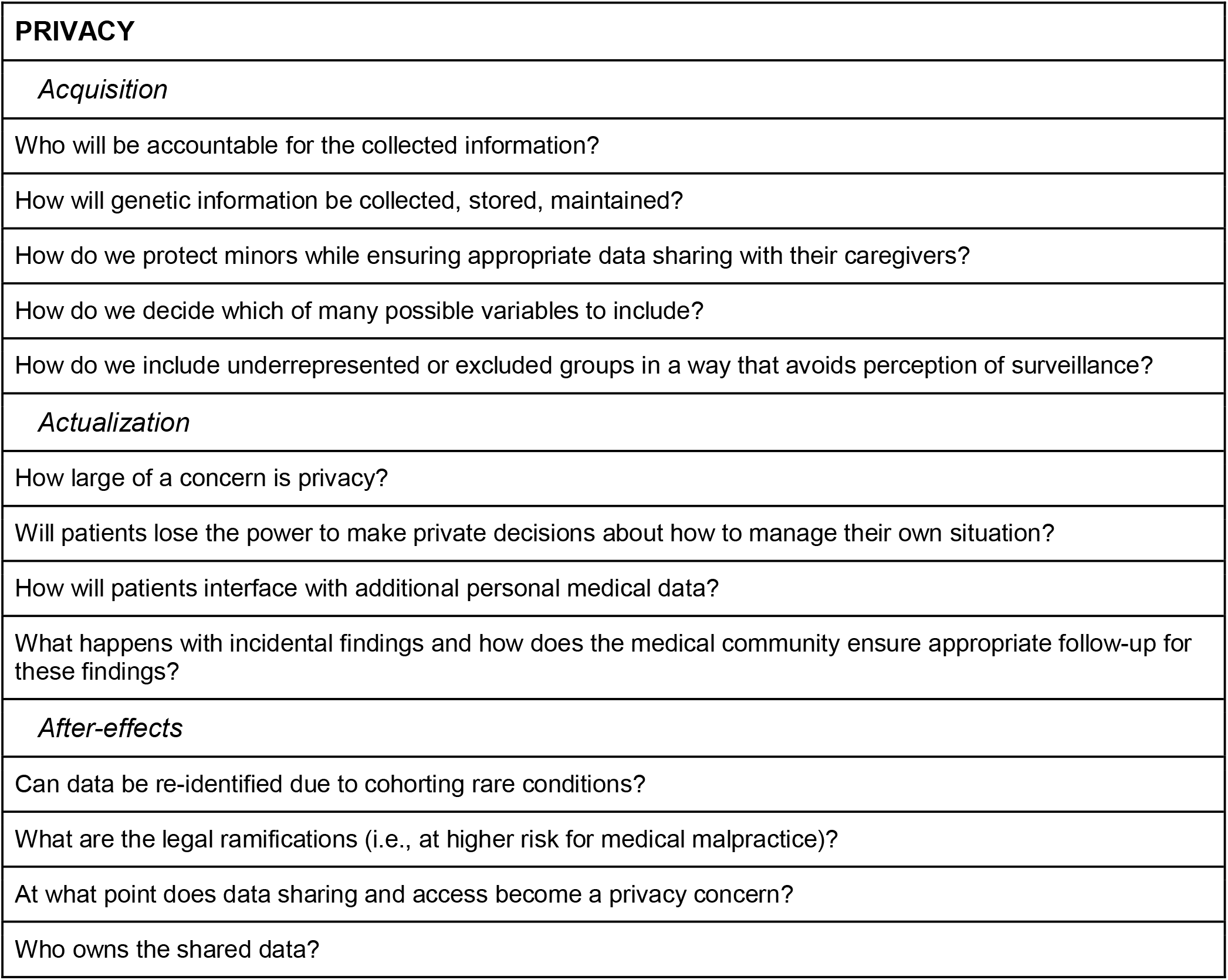
Participant concerns regarding privacy in the precision medicine process.

|  |
| --- |
| **PRIVACY** |
| *Acquisition* |
| Who will be accountable for the collected information? |
| How will genetic information be collected, stored, maintained? |
| How do we protect minors while ensuring appropriate data sharing with their caregivers? |
| How do we decide which of many possible variables to include? |
| How do we include underrepresented or excluded groups in a way that avoids perception of surveillance? |
| *Actualization* |
| How large of a concern is privacy? |
| Will patients lose the power to make private decisions about how to manage their own situation? |
| How will patients interface with additional personal medical data? |
| What happens with incidental findings and how does the medical community ensure appropriate follow-up for these findings? |
| *After-effects* |
| Can data be re-identified due to cohorting rare conditions? |
| What are the legal ramifications (i.e., at higher risk for medical malpractice)? |
| At what point does data sharing and access become a privacy concern? |
| Who owns the shared data? |

**Table 3:**
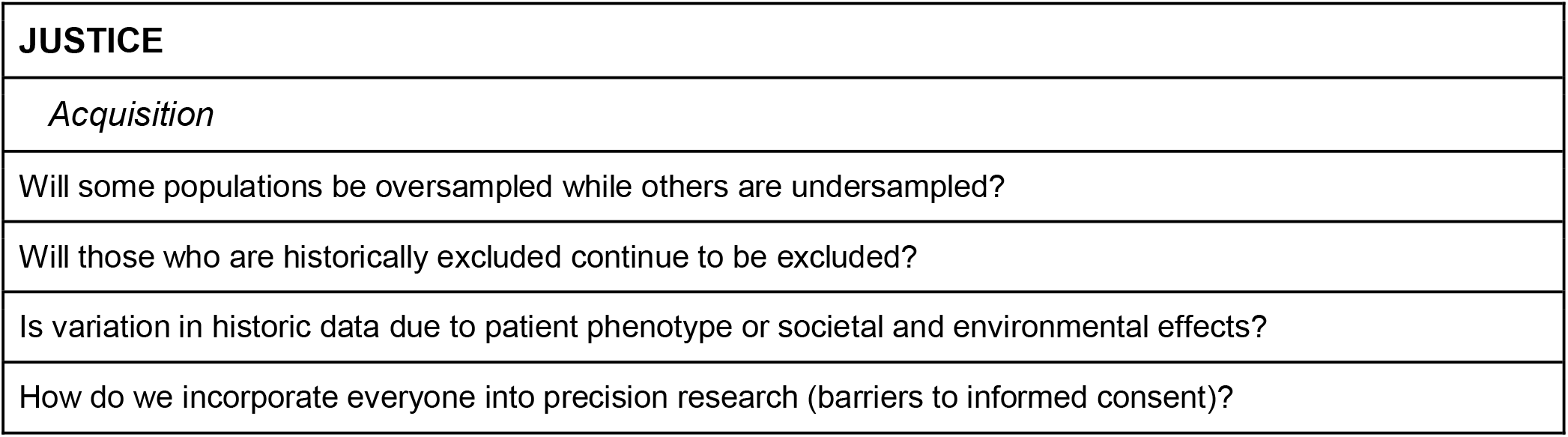
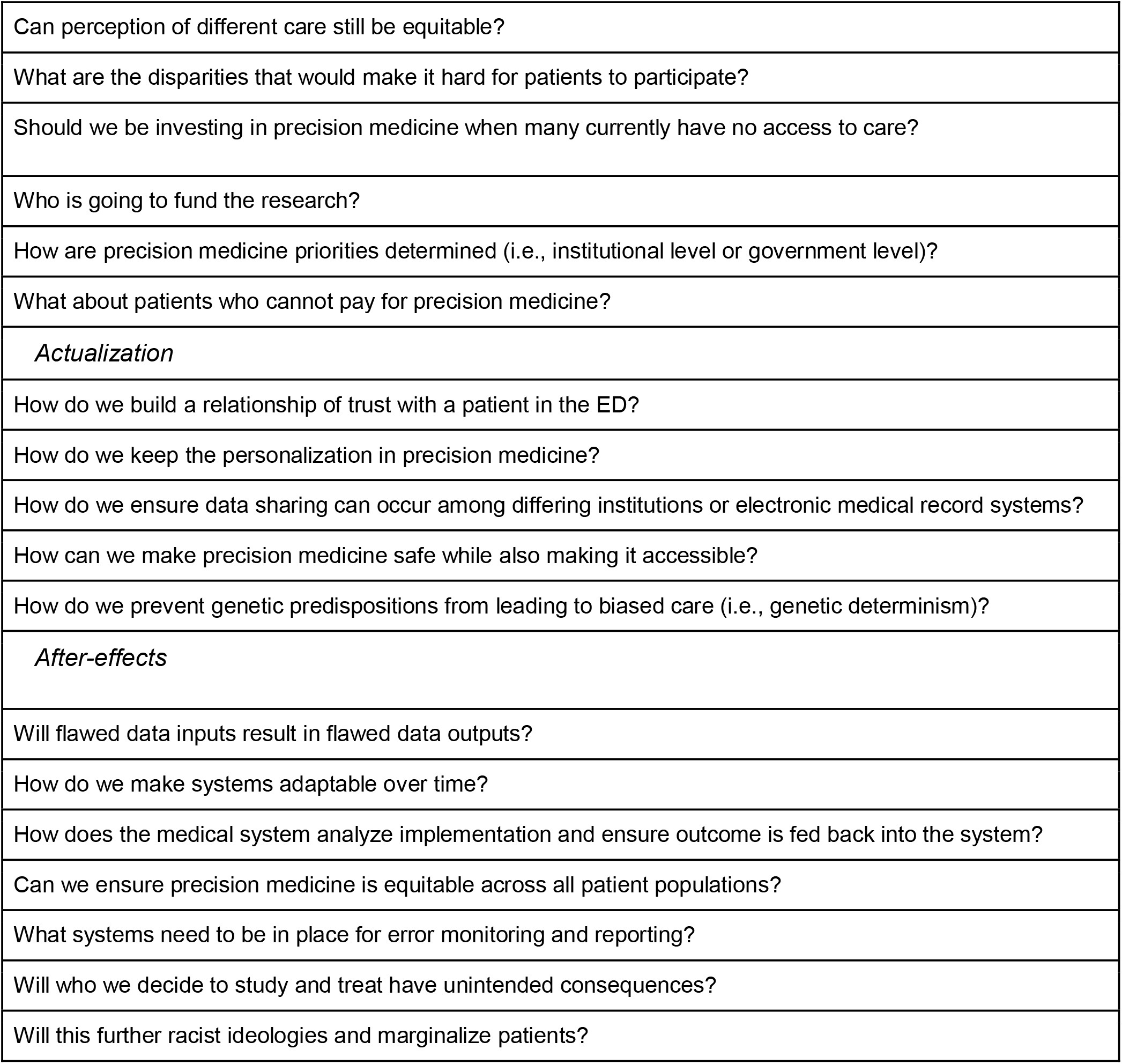
Participant concerns about how to ensure just care in the precision medicine model and process.

**Figure 2:**
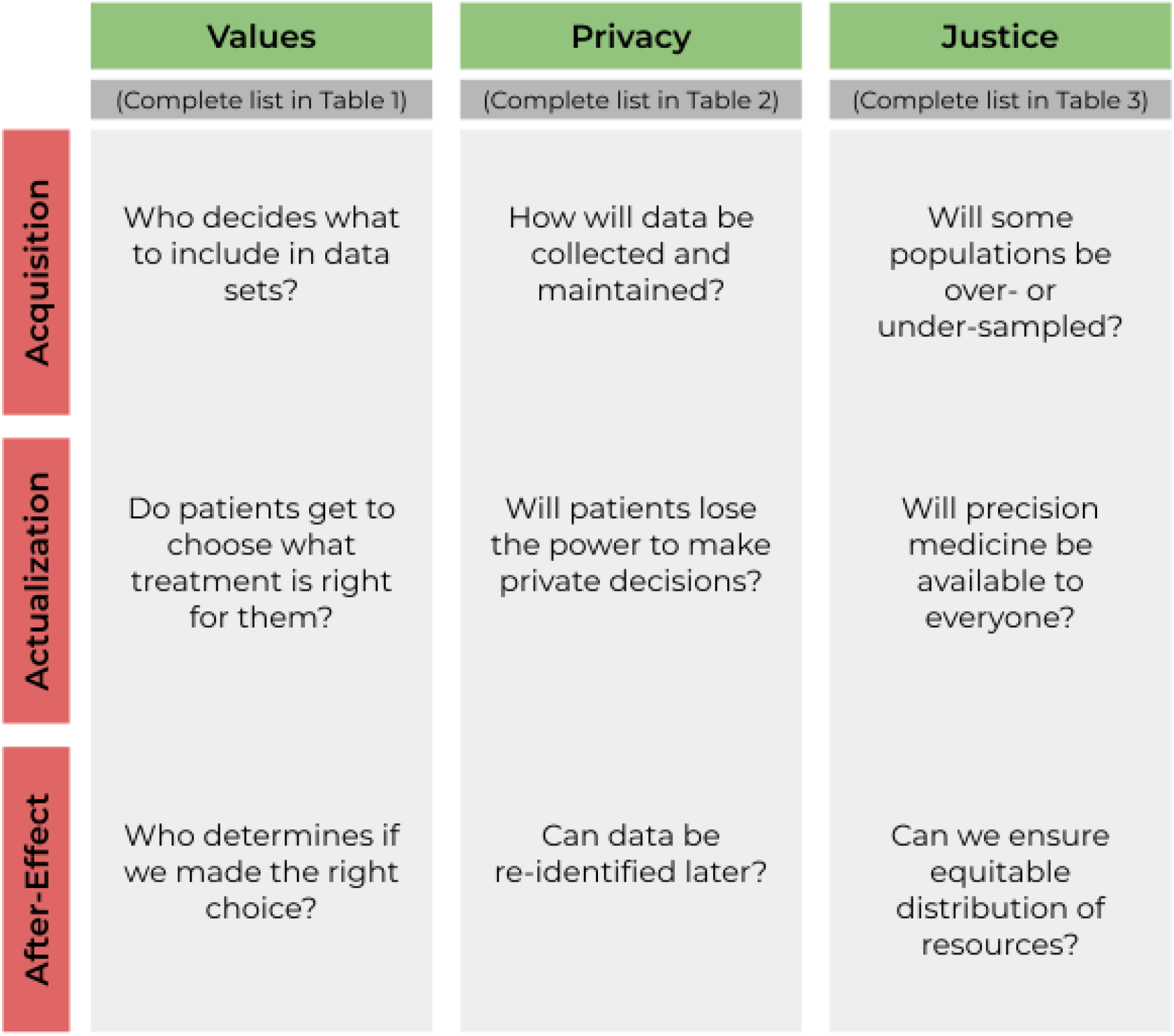
Priority matrix of ethical concerns for precision medicine.

## Discussion

Precision medicine research and practice in the emergency medicine context requires attention to the specific ethical concerns of those working in this space in order to avoid potential pitfalls and promote inclusion and equity. Working with a diverse group of emergency physicians, we used a modified NGT approach to identify critical areas in need of research at the intersection of emergency medicine, precision medicine, and ethics. The resulting consensus framework highlights critical areas of focus for precision medicine research in emergency medicine that are cognizant of the limits and challenges of data acquisition, the actualization of precision medicine in practice, and the potential after-effects of this pursuit for our patients and communities. Three major themes emerged from the modified NGT: values, privacy, and justice.

### Shared Ethical Concerns: Values, Privacy, Justice

A recurring theme in discussion was “whose values are we accounting for?” Participants acknowledged the sometimes conflicting needs of the individual versus those of the system. Providers may recommend medications which prolong survival, but side effects may be undesirable or the cost prohibitive for some patients.^38^ The definition of high quality care envisioned by precision medicine may not align with the individual goals of patients. There were many questions raised regarding challenges to patient autonomy and to the ambiguity of who is the ultimate decision maker in precision medicine–and how that might change over time, as the needs of the many may be at odds with the needs of a few. The ethical principle of autonomy is considered perhaps the most important principle in medical ethics.^39^ Respect for autonomy requires that patients are able to make their own decisions, incorporating their own values, but the limits of patient autonomy remain under debate.^40^

Ensuring that patient autonomy is maintained can be particularly challenging in the emergency care context. In the emergency department, providers make decisions with patients in a time-constrained setting, particularly compared to those historical clinical environments of precision medicine, such as oncology. Not only does this limit time for discussion, it may also create a sense of external pressure to make a decision.^41^ Might other competing factors in this clinical setting also ultimately limit patient and physician choice given precision medicine’s reliance on optimizing measurable and scalable outcomes? The idea of autonomy becomes less clear when it comes to data at the population level: whose value system will be used to make decisions once data has moved from the individual care setting to community responsibilities? Will patient values (e.g., best outcome) align with broader system values (e.g., cost-effectiveness) when considering precision medicine research and recommendations? To what extent might patients ultimately lose elements of individual autonomy once their data is incorporated into communal data?

This latter concept underscores the importance of privacy in precision medicine. Confidentiality, a core ethical principle of emergency medicine,^42^ requires providers ensure their patient’s privacy. The practice of precision medicine in the emergency department will likely require massive amounts of data transiting across institutions and from personal devices. Ensuring the safety and security of this protected information is a major hurdle for precision medicine initiatives, and the above-mentioned time-sensitive nature of the emergency department encounter makes it necessary to share this data in real-time, which is unique from other medical specialties.^43–46^ Additionally, when patients know that the information they provide to the system will be used to make recommendations for themselves or others, it may have several effects. First, patients may not accurately disclose their values and personal concerns if they do not have trust in the safety of their information and its use. For example, alcohol use and history for the allotment of liver transplant is one area which may leave patients in a moral quandary, unsure whether to be open with their behavior but fearing the effects of those disclosures.^47,48^ Furthermore, patients may be concerned that their personal choices may have adverse effects on people like them, providing another ethical dilemma for how and what to disclose. Privacy includes not only the protection of one’s identity and personal information, but also the ability to make choices without interference.^49^ Perceptions of trustworthiness will impact the success of any scientific or research endeavor, particularly among historically disadvantaged populations.^50^

But perhaps nowhere is the complex tension between values and privacy felt more than in the realm of justice. A commitment to justice is in the historical roots of emergency medicine and it is no surprise that our consensus framework includes this as a key area for research in precision medicine as it grows in the emergency department and in the prehospital setting.^51–53^ Emergency physicians carry the distinct duty under federal law to provide care to anyone who presents for emergency care.^54^ This duty is widely recognized to rest on a fundamental moral responsibility to provide care “promptly and expertly, without prejudice or partiality”.^42^ At first it may seem that precision medicine would help to limit personal biases which might affect care, using data as opposed to gestalt or local practice variation. But structural biases may encode results and outcomes on patients that might inadvertently limit the benefits of the precision medicine model. Obvious examples include the use of race-adjusted creatinine to allocate critical resources which has occurred for years and affects the validity of results used to make targeted recommendations.^55^

Justice, or more specifically with respect to medical ethics, distributive justice, emphasizes that resources are to be distributed fairly, equitably, and appropriately. This can be heavily hindered if providers rely on precision models to make recommendations without a critical appraisal of how well the results fit the patient in front of them. Even if done well, the outputs of precision medicine may not be available to all.^56^ Precision medicine which is ultimately only accessible to a few but built on the data of many, would violate the principle of justice and would be an affront to the growing field of social emergency medicine. In oncology, an area with a much longer history of precision medicine, 78% of patients and providers reported concerns regarding the accessibility of precision medicine.^57^

### Cross-Cutting Themes & the Process of Precision Medicine

Participant discussion emphasized the need to investigate concerns of justice, privacy, and values from a longitudinal perspective: data acquisition, actualization, and application after-effects. In doing so, additional cross cutting themes emerged (Supplemental Table 1), many of which are shared with other applications of precision medicine.

#### Data Availability and Acquisition

Ultimately, the trust patients and providers have in the recommendations of precision medicine begins with the data that are used and collected. In emergency medicine, we, as providers, stand prepared to address a range of conditions for any patient, at any time. However, we often cannot rely on complete patient histories (e.g., due to the severity of illness or mental status changes frequently encountered in emergency medicine) or access to prior medical records (e.g., our patients may be uninsured, underinsured, and without access to routine primary care providers.^58^ Precision medicine approaches will thus need to be deployable in a timely manner and require only what limited patient-specific information is available to emergency physicians.

Furthermore, even when data are available, precision medicine approaches may serve only to perpetuate systemic flaws in the healthcare system if the data used is inaccurate, biased, or exclusionary from the start.^7,59^ Data may reflect bias with respect to inclusion^60^ as well as bias stemming from structural racism. A 12-year review of the National Hospital Ambulatory Medical Survey (NHAMCS) showed that Black patients in the emergency department were 10% less likely to receive immediate or emergent Emergency Severity Index (ESI) scores compared to white patients, 10% less likely than white patients to be admitted, but 1.26 more likely to die in the emergency department or the hospital than white patients.^58^ If these data were used to make recommendations about who and where to direct resources, it may serve only to further amplify the pre-existing failures to address and reduce bias that are present in the data and its acquisition.

#### Choice

Beyond the issue of timing, there are additional logistic concerns for the actualization of precision medicine in emergency departments, including costs, legal issues and patient education. Efforts made to collate data from emergency care settings to be used in precision medicine research may result in products or treatment pathways unaffordable to these patients.^61–63^ This may be mitigated through insurers,^64^ but the emergency department is a safety net, and as such this approach may not benefit the most vulnerable patients while those who are insured may find that their plans have little incentive to offer this option.^65^ Research has shown that the cost of precision medicine is a concern shared by patients and providers alike.^56,57^

#### Informed Decision Making

Even when payment is not an issue, a data- and machine-supported approach may not be conducive to truly informed consent given the limitations of adequately educating emergency department patients regarding how precision medicine and its associated research works.^56,66^ Beyond the challenge of consenting in the emergency context, providers must also consider issues regarding disclosing incidental information and the potential changes associated with the use of data over time which interconnected to other growing datasets. Stewardship of these data, defined as “the responsible management of something entrusted to one’s care”,^67^ will be critical to maintaining trust of patients and communities as this science evolves.^50^

#### Avoiding Reductionism

Despite the promise of precision medicine to deliver individually tailored care, it, like traditional approaches, requires that patients be categorized into discrete groups by some shared characteristics. The choice of categories and the act of assigning patients to these groups has its own meaning and implications.^4^

Discussion in this NGT process highlighted the need to actively maintain a holistic view of the patient without being overly reductive to these categories. Currently, only a limited amount of data are collected with respect to health measures or lab studies, and even less is recorded for social determinants of health. Future work is needed to better incorporate environmental and social factors into precision medicine models to better delineate the contribution of these critical factors to emergency medicine presentations, outcomes, and recommendations for care.

### Limitations

While NGT aims to limit the biases associated with other consensus models, expertise bias is still a potential limitation of this study, given that all participants were aware of and knowledgeable on this topic beforehand. However, we took several steps to limit its impact. NGT, by way of its structured format, is meant to limit the influence of any one dominant individual in discussion.^37^ Further, we included a diverse spectrum of emergency physicians from residents to attending physicians with varying years of experience, as well as emergency physicians with a variety of subspeciality training. While the number of participants enrolled is in the ideal range for an NGT,^24^ nevertheless, the number of participants is small and future research could benefit from additional strategies that would incorporate a higher number of emergency physicians and experts from outside of the field which might offer perspective to our work.

### Conclusions

In summary, realizing the promise of precision medicine’s “right person, right place, right time” must be balanced by the context of the “anyone, anywhere, any time” of emergency medicine. Our work has illuminated a range of concerns for the development of precision emergency medicine, and provides a focused framework for the most pressing ethical considerations along the continuum of data acquisition to implementation. This framework may be used to direct research and innovation toward addressing these challenges in the emergency medicine setting.

## Data Availability

All data produced in the present work are contained in the manuscript.

---

**Appendix Table 1:**
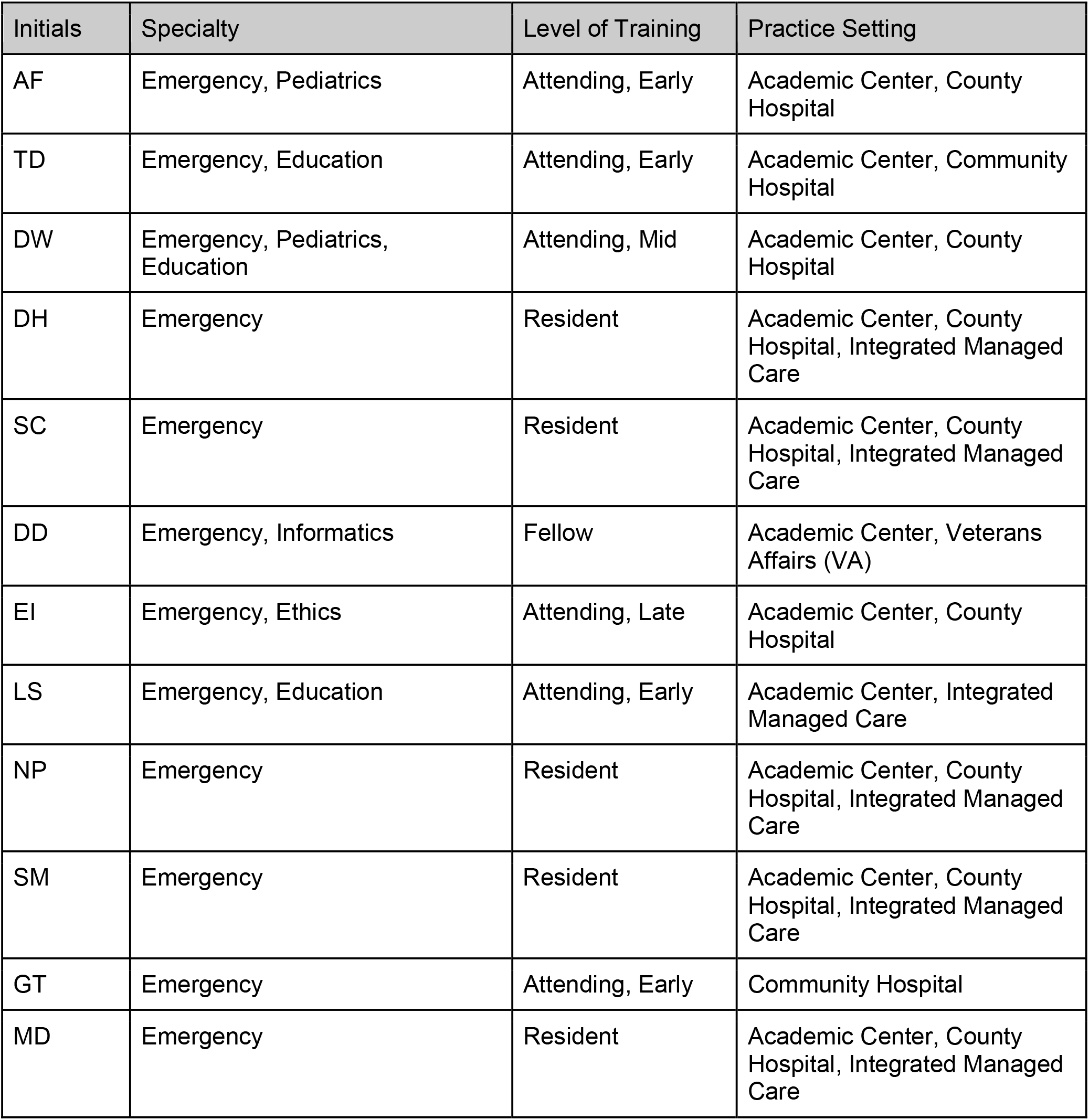
Nominal Group participant specialty and level of training.

**Appendix Table 2:**
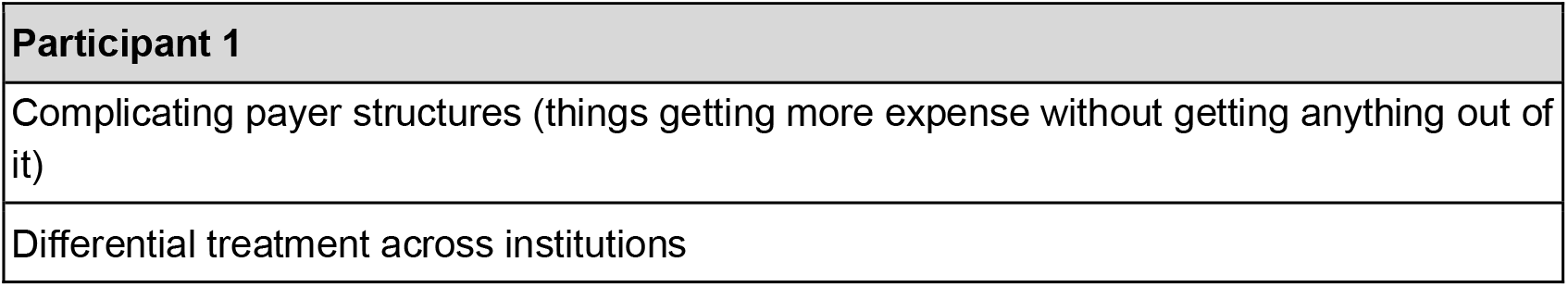
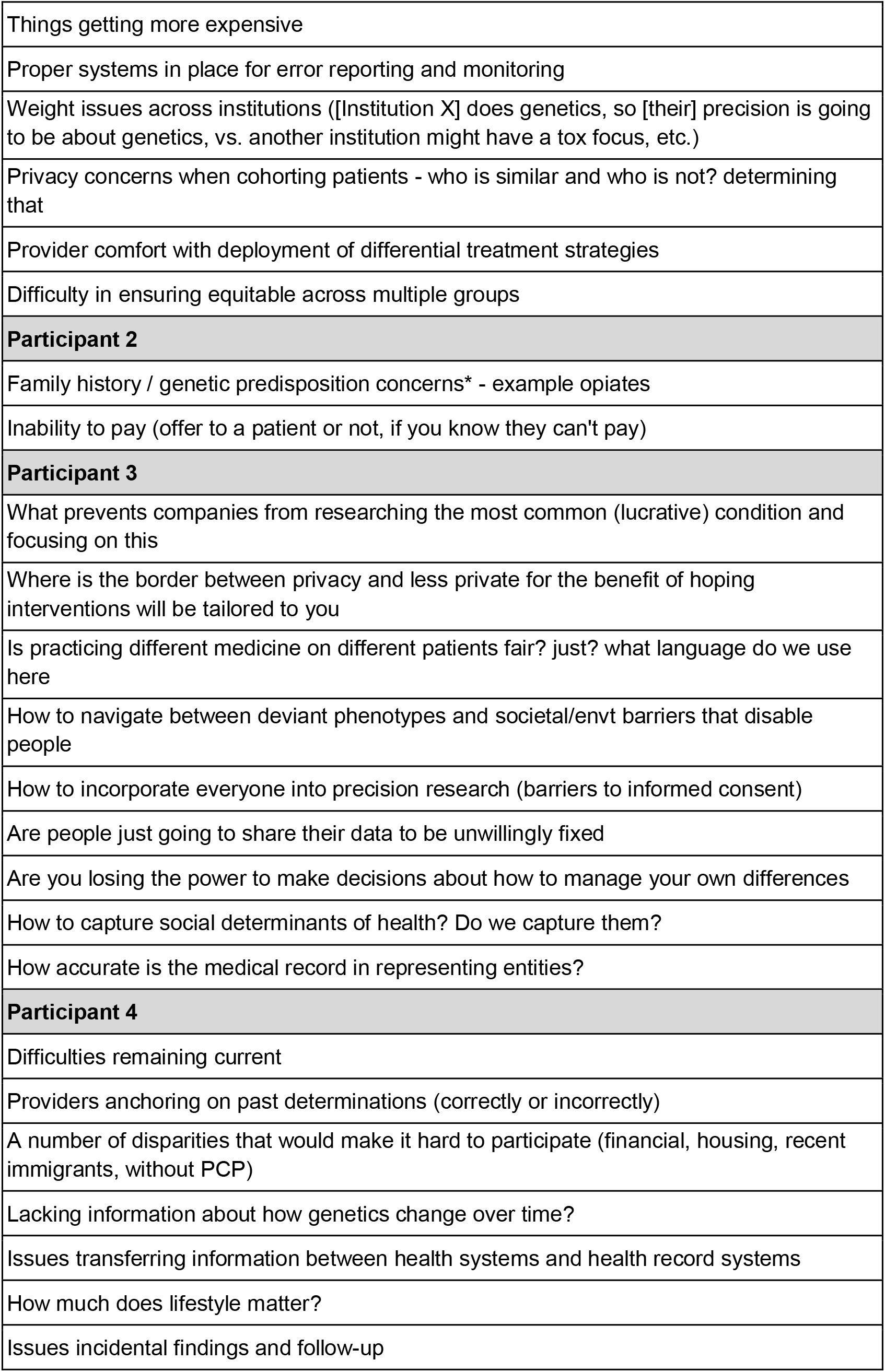
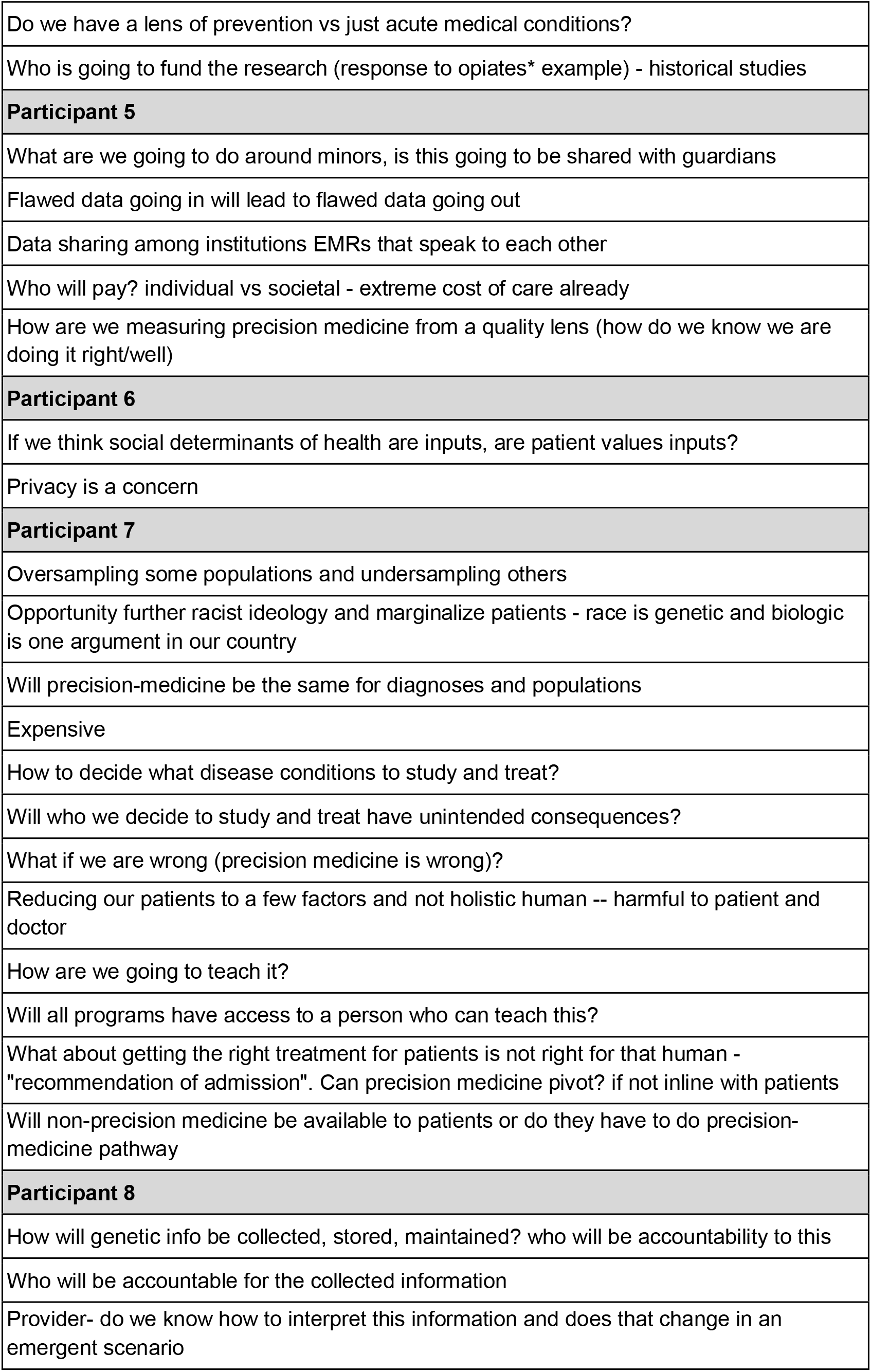
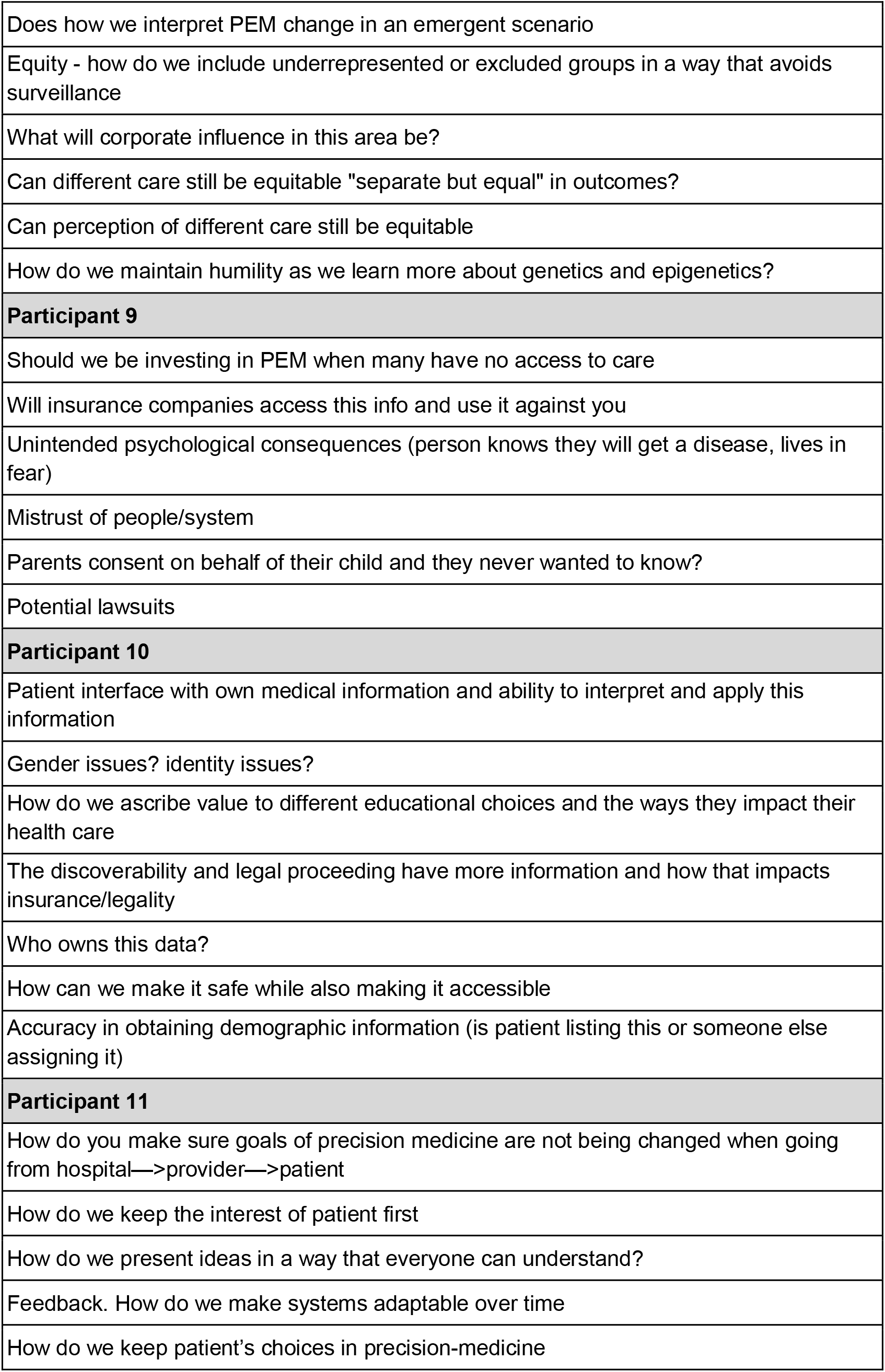
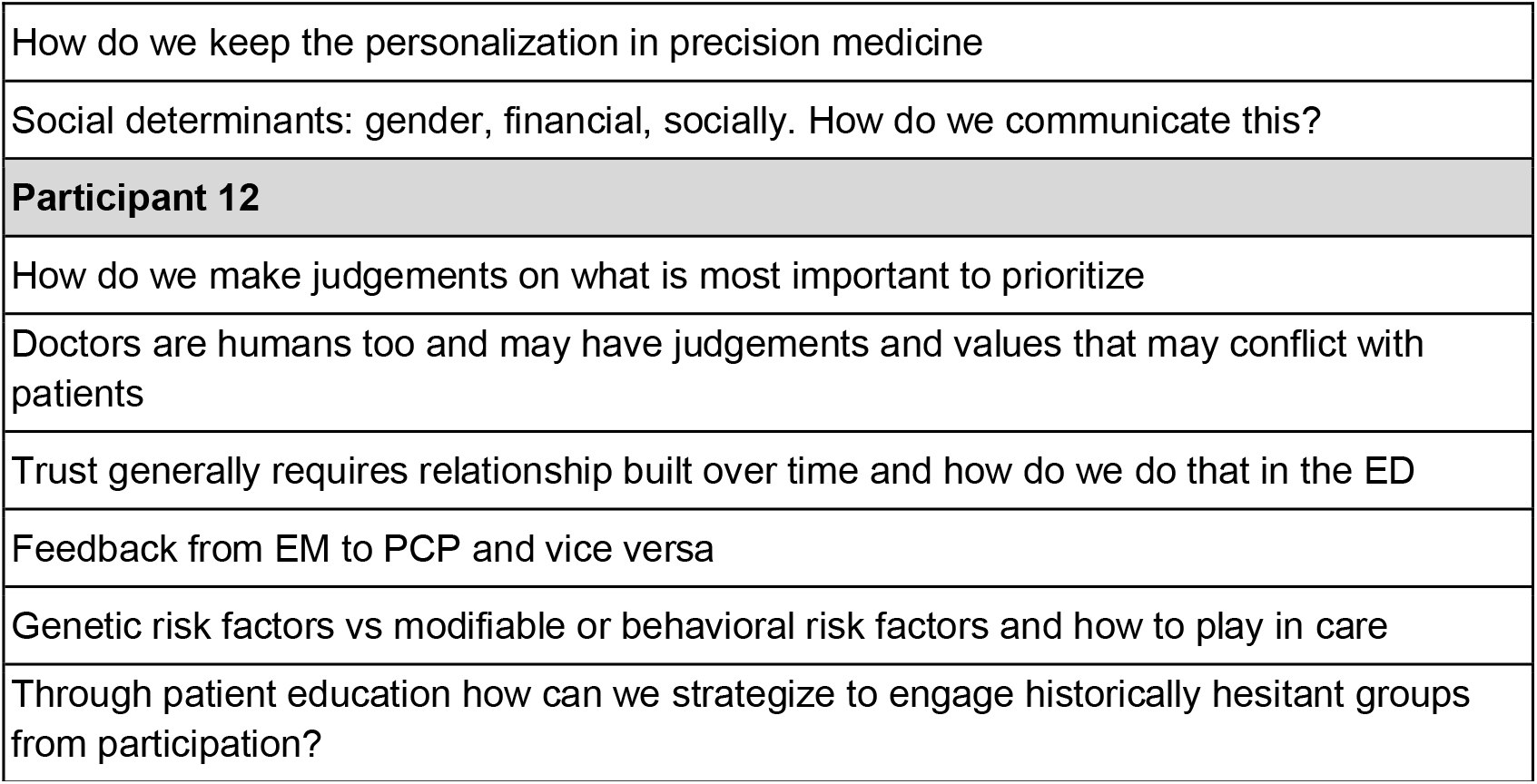
Complete list of questions generated by participants in steps one and two.

**Appendix Table 1:**
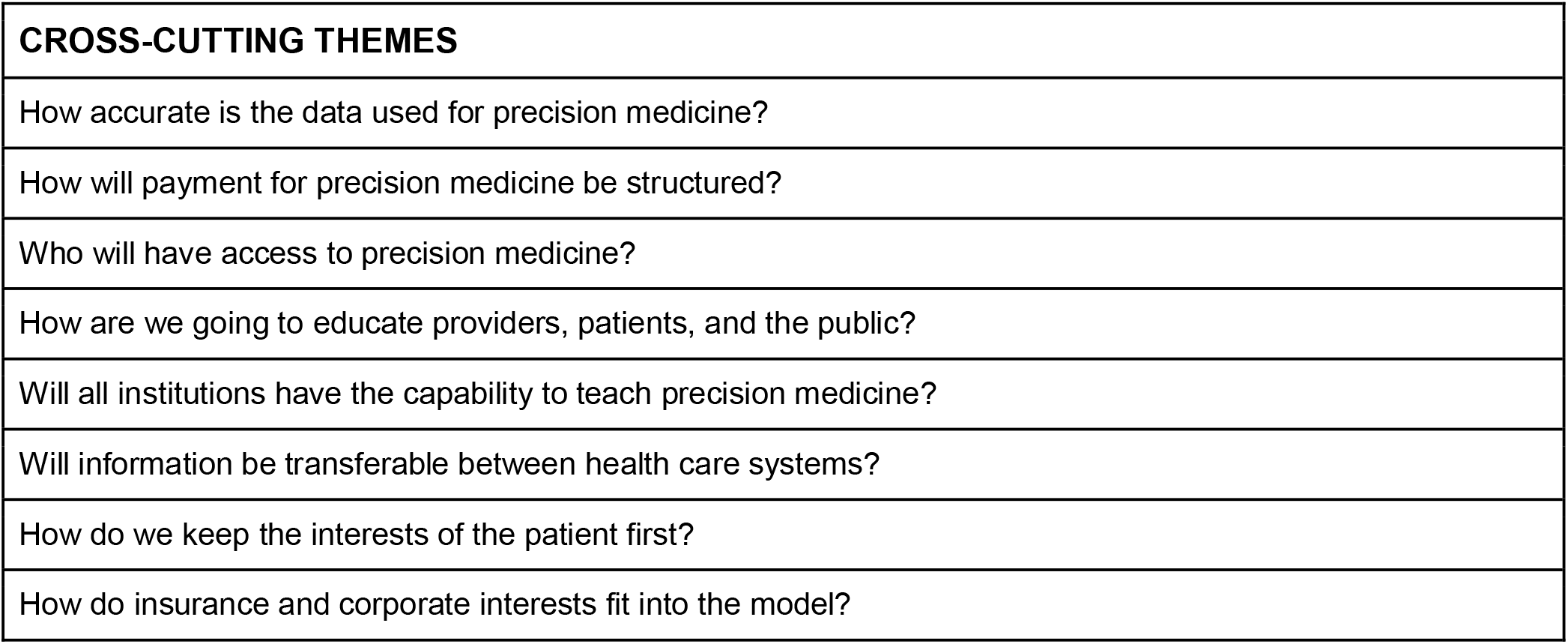
Cross-Cutting Themes

## References

1. Salari P, Larijani B. Ethical Issues Surrounding Personalized Medicine: A Literature Review. Acta Med Iran. 2017;55(3):209–217.

2. Liu X, Luo X, Jiang C, Zhao H. Difficulties and challenges in the development of precision medicine. Clin Genet. 2019;95(5):569–574.

3. Sitapati A, Kim H, Berkovich B, et al. Integrated precision medicine: the role of electronic health records in delivering personalized treatment. Wiley Interdiscip Rev Syst Biol Med. 2017;9(3). doi:10.1002/wsbm.1378

4. Seymour CW, Gomez H, Chang CCH, et al. Precision medicine for all? Challenges and opportunities for a precision medicine approach to critical illness. Crit Care. 2017;21(1):257.

5. Mensah GA, Jaquish C, Srinivas P, et al. Emerging Concepts in Precision Medicine and Cardiovascular Diseases in Racial and Ethnic Minority Populations. Circ Res. 2019;125(1):7–13.

6. Lee SSJ, Fullerton SM, Saperstein A, Shim JK. Ethics of inclusion: Cultivate trust in precision medicine. Science. 2019;364(6444):941–942.

7. Geneviève LD, Martani A, Shaw D, Elger BS, Wangmo T. Structural racism in precision medicine: leaving no one behind. BMC Med Ethics. 2020;21(1):17.

8. Stein JN, Charlot M, Cykert S. Building Toward Antiracist Cancer Research and Practice: The Case of Precision Medicine. JCO Oncol Pract. 2021;17(5):273–277.

9. Goetz LH, Schork NJ. Personalized medicine: motivation, challenges, and progress. Fertil Steril. 2018;109(6):952–963.

10. Elemento O. The future of precision medicine: towards a more predictive personalized medicine. Emerg Top Life Sci. 2020;4(2):175–177.

11. Hill-Briggs F, Adler NE, Berkowitz SA, et al. Social Determinants of Health and Diabetes: A Scientific Review. Diabetes Care. Published online November 2, 2020. doi:10.2337/dci20-0053

12. Marmot M, Allen JJ. Social determinants of health equity. Am J Public Health. 2014;104 Suppl 4:S517–S519.

13. Kalra D. The importance of real-world data to precision medicine. Per Med. 2019;16(2):79–82.

14. Hollister B, Bonham VL. Should Electronic Health Record-Derived Social and Behavioral Data Be Used in Precision Medicine Research? AMA J Ethics. 2018;20(9):E873–E880.

15. Sukumar SR, Natarajan R, Ferrell RK. Quality of Big Data in health care. Int J Health Care Qual Assur. 2015;28(6):621–634.

16. Desharnais S. Current Uses of Large Data Sets to Assess the Quality of Providers: Construction of Risk-Adjusted Indexes of Hospital Performance. Int J Technol Assess Health Care. 1990;6(2):229–238.

17. Williams WW, Hogan JW, Ingelfinger JR. Time to Eliminate Health Care Disparities in the Estimation of Kidney Function. N Engl J Med. 2021;385(19):1804–1806.

18. Wiens J, Saria S, Sendak M, et al. Do no harm: a roadmap for responsible machine learning for health care. Nat Med. 2019;25(9):1337–1340.

19. Char DS, Shah NH, Magnus D. Implementing Machine Learning in Health Care — Addressing Ethical Challenges. New England Journal of Medicine. 2018;378(11):981–983. doi:10.1056/nejmp1714229

20. Owens K, Walker A. Those designing healthcare algorithms must become actively anti-racist. Nat Med. 2020;26(9):1327–1328.

21. Rajkomar A, Hardt M, Howell MD, Corrado G, Chin MH. Ensuring Fairness in Machine Learning to Advance Health Equity. Ann Intern Med. 2018;169(12):866–872.

22. Marcozzi D, Carr B, Liferidge A, Baehr N, Browne B. Trends in the Contribution of Emergency Departments to the Provision of Hospital-Associated Health Care in the USA. Int J Health Serv. 2018;48(2):267–288.

23. Hsia RY, Sabbagh SH, Guo J, Nuckton TJ, Niedzwiecki MJ. Trends in the utilisation of emergency departments in California, 2005–2015: a retrospective analysis. BMJ Open. 2018;8(7):e021392.

24. McMillan SS, King M, Tully MP. How to use the nominal group and Delphi techniques. Int J Clin Pharm. 2016;38(3):655–662.

25. Ranney ML, Fletcher J, Alter H, et al. A Consensus-Driven Agenda for Emergency Medicine Firearm Injury Prevention Research. Ann Emerg Med. 2017;69(2):227–240.

26. Elder E, Johnston AN, Byrne JH, Wallis M, Crilly J. Core components of a staff wellness strategy in emergency departments: A clinician-informed nominal group study. Emerg Med Australas. 2021;33(1):25–33.

27. Nik Hisamuddin R, Tuan Hairulnizam TK. Developing Key Performance Indicators for Emergency Department of Teaching Hospitals: A Mixed Fuzzy Delphi and Nominal Group Technique Approach. Malays J Med Sci. 2022;29(2):114–125.

28. Peltzer-Jones J, Nordstrom K, Currier G, Berlin JS, Singh C, Schneider S. A Research Agenda for Assessment and Management of Psychosis in Emergency Department Patients. West J Emerg Med. 2019;20(2):403–408.

29. Members of the SAEM Consensus Conference Emergency Medical Services Subcommittee, Adelgais KM, Hansen M, et al. Establishing the key outcomes for pediatric emergency medical services research. Acad Emerg Med. 2018;25(12):1345–1354.

30. Marynowski-Traczyk D, Wallis M, Broadbent M, et al. Optimising emergency department and acute care for people experiencing mental health problems: a nominal group study. Aust Health Rev. 2022;46(5):519–528.

31. Safdar B, Greenberg MR. Organization, execution and evaluation of the 2014 Academic Emergency Medicine consensus conference on Gender-Specific Research in Emergency Care - an executive summary. Acad Emerg Med. 2014;21(12):1307–1317.

32. Harvey N, Holmes CA. Nominal group technique: an effective method for obtaining group consensus. Int J Nurs Pract. 2012;18(2):188–194.

33. Kottmann A, Krüger AJ, Sunde GA, et al. Establishing quality indicators for pre-hospital advanced airway management: a modified nominal group technique consensus process. Br J Anaesth. 2022;128(2):e143–e150.

34. Sunde GA, Kottmann A, Heltne JK, et al. Standardised data reporting from pre-hospital advanced airway management - a nominal group technique update of the Utstein-style airway template. Scand J Trauma Resusc Emerg Med. 2018;26(1):46.

35. Nelson V, Dubov A, Morton K, Fraenkel L. Using nominal group technique among resident physicians to identify key attributes of a burnout prevention program. PLoS One. 2022;17(3):e0264921.

36. Gallagher M, Hares T, Spencer J, Bradshaw C, Webb I. The nominal group technique: a research tool for general practice? Fam Pract. 1993;10(1):76–81.

37. Humphrey-Murto S, Varpio L, Gonsalves C, Wood TJ. Using consensus group methods such as Delphi and Nominal Group in medical education research. Med Teach. 2017;39(1):14–19.

38. Kelly TD, Hawk KF, Samuels EA, Strayer RJ, Hoppe JA. Improving Uptake of Emergency Department-initiated Buprenorphine: Barriers and Solutions. West J Emerg Med. 2022;23(4):461–467.

39. Sedig L. What’s the Role of Autonomy in Patient- and Family-Centered Care When Patients and Family Members Don’t Agree? AMA Journal of Ethics. 2016;18(1):12–17.

40. Entwistle VA, Carter SM, Cribb A, McCaffery K. Supporting patient autonomy: the importance of clinician-patient relationships. J Gen Intern Med. 2010;25(7):741–745.

41. Naess AC, Foerde R, Steen PA. Patient autonomy in emergency medicine. Med Health Care Philos. 2001;4(1):71–77.

42. American College of Emergency Physicians. Code of ethics for emergency physicians. Ann Emerg Med. 2008;52(5):581–590.

43. Lee SSJ, Cho MK, Kraft SA, et al. “I don’t want to be Henrietta Lacks”: diverse patient perspectives on donating biospecimens for precision medicine research. Genet Med. 2018;21(1):107–113.

44. Ratcliff CL, Kaphingst KA, Jensen JD. When Personal Feels Invasive: Foreseeing Challenges in Precision Medicine Communication. J Health Commun. 2018;23(2):144–152.

45. Stiles D, Appelbaum PS. Cases in Precision Medicine: Concerns About Privacy and Discrimination After Genomic Sequencing. Ann Intern Med. 2019;170(10):717–721.

46. Sankar PL, Parker LS. The Precision Medicine Initiative’s All of Us Research Program: an agenda for research on its ethical, legal, and social issues. Genet Med. 2016;19(7):743–750.

47. Moss AH, Siegler M. Should alcoholics compete equally for liver transplantation? JAMA. 1991;265(10):1295–1298.

48. Sedki M, Ahmed A, Goel A. Ethical and allocation issues in liver transplant candidates with alcohol related liver disease. Transl Gastroenterol Hepatol. 2022;7:26.

49. Geiderman JM, Moskop JC, Derse AR. Privacy and confidentiality in emergency medicine: obligations and challenges. Emerg Med Clin North Am. 2006;24(3):633–656.

50. Kraft SA, Cho MK, Gillespie K, et al. Beyond Consent: Building Trusting Relationships With Diverse Populations in Precision Medicine Research. Am J Bioeth. 2018;18(4):3–20.

51. Zink BJ. Social justice, egalitarianism, and the history of emergency medicine. Virtual Mentor. 2010;12(6):492–494.

52. Anderson ES, Lippert S, Newberry J, Bernstein E, Alter HJ, Wang NE. Addressing Social Determinants of Health from the Emergency Department through Social Emergency Medicine. West J Emerg Med. 2016;17(4):487–489.

53. Ben-Assuli O. Electronic health records, adoption, quality of care, legal and privacy issues and their implementation in emergency departments. Health Policy. 2015;119(3):287–297.

54. Brown HL, Brown TB. EMTALA: The Evolution of Emergency Care in the United States. J Emerg Nurs. 2019;45(4):411–414.

55. Boulware LE, Purnell TS, Mohottige D. Systemic Kidney Transplant Inequities for Black Individuals: Examining the Contribution of Racialized Kidney Function Estimating Equations. JAMA Netw Open. 2021;4(1):e2034630.

56. Erdmann A, Rehmann-Sutter C, Bozzaro C. Patients’ and professionals’ views related to ethical issues in precision medicine: a mixed research synthesis. BMC Med Ethics. 2021;22(1):116.

57. Miller AM, Garfield S, Woodman RC. Patient and provider readiness for personalized medicine. Pers Med Oncol.

58. Zhang X, Carabello M, Hill T, Bell SA, Stephenson R, Mahajan P. Trends of Racial/Ethnic Differences in Emergency Department Care Outcomes Among Adults in the United States From 2005 to 2016. Front Med. 2020;7:300.

59. Mersha TB, Abebe T. Self-reported race/ethnicity in the age of genomic research: its potential impact on understanding health disparities. Hum Genomics. 2015;9:1.

60. Manrai AK, Patel CJ, Ioannidis JPA. In the Era of Precision Medicine and Big Data, Who Is Normal? JAMA. 2018;319(19):1981–1982.

61. O’Sullivan BP, Orenstein DM, Milla CE. Pricing for orphan drugs: will the market bear what society cannot? JAMA. 2013;310(13):1343–1344.

62. Ferkol T, Quinton P. Precision Medicine: At What Price? Am J Respir Crit Care Med. 2015;192(6):658–659.

63. Ramaswami R, Bayer R, Galea S. Precision Medicine from a Public Health Perspective. Annu Rev Public Health. 2018;39:153–168.

64. Geruso M, Jena AB, Layton TJ. Will Personalized Medicine Mean Higher Costs for Consumers? Harvard Business Review. Published online March 1, 2018. Accessed October 27, 2022. https://hbr.org/2018/03/will-personalized-medicine-mean-higher-costs-for-consumers

65. Horgan D, Jansen M, Leyens L, et al. An index of barriers for the implementation of personalised medicine and pharmacogenomics in Europe. Public Health Genomics. 2014;17(5-6):287–298.

66. Fiore RN, Goodman KW. Precision medicine ethics: selected issues and developments in next-generation sequencing, clinical oncology, and ethics. Curr Opin Oncol. 2016;28(1):83–87.

67. Fullerton SM, Anderson NR, Guzauskas G, Freeman D, Fryer-Edwards K. Meeting the governance challenges of next-generation biorepository research. Sci Transl Med. 2010;2(15):15cm3.

