## Supplementary material for "Identifying High Priority Ethical Challenges for Precision Emergency Medicine - A Nominal Group Study": S1

Table S1: Nominal Group participant specialty and level of training.

| Initials | Specialty | Level of Training | Practice Setting |
| --- | --- | --- | --- |
| AF | Emergency, Pediatrics | Attending, Early | Academic Center, County Hospital |
| TD | Emergency, Education | Attending, Early | Academic Center, Community Hospital |
| DW | Emergency, Pediatrics, Education | Attending, Mid | Academic Center, County Hospital |
| DH | Emergency | Resident | Academic Center, County Hospital, Integrated Managed Care |
| SC | Emergency | Resident | Academic Center, County Hospital, Integrated Managed Care |
| DD | Emergency, Informatics | Fellow | Academic Center, Veterans Affairs (VA) |
| EI | Emergency, Ethics | Attending, Late | Academic Center, County Hospital |
| LS | Emergency, Education | Attending, Early | Academic Center, Integrated Managed Care |
| NP | Emergency | Resident | Academic Center, County Hospital, Integrated Managed Care |
| SM | Emergency | Resident | Academic Center, County Hospital, Integrated Managed Care |
| GT | Emergency | Attending, Early | Community Hospital |
| MD | Emergency | Resident | Academic Center, County Hospital, Integrated Managed Care |

Table S2: Complete list of questions generated by participants in steps one and two.

**Participant 1**

Complicating payer structures (things getting more expense without getting anything out of it)

Differential treatment across institutions

Things getting more expensive

Proper systems in place for error reporting and monitoring

Weight issues across institutions ([Institution X] does genetics, so [their] precision is going to be about genetics, vs. another institution might have a tox focus, etc.)

Privacy concerns when cohorting patients - who is similar and who is not? determining that

Provider comfort with deployment of differential treatment strategies

Difficulty in ensuring equitable across multiple groups

**Participant 2**

Family history / genetic predisposition concerns* - example opiates

Inability to pay (offer to a patient or not, if you know they can't pay)

**Participant 3**

What prevents companies from researching the most common (lucrative) condition and focusing on this

Where is the border between privacy and less private for the benefit of hoping interventions will be tailored to you

Is practicing different medicine on different patients fair? just? what language do we use here

How to navigate between deviant phenotypes and societal/envt barriers that disable people

How to incorporate everyone into precision research (barriers to informed consent)

Are people just going to share their data to be unwillingly fixed

Are you losing the power to make decisions about how to manage your own differences

How to capture social determinants of health? Do we capture them?

How accurate is the medical record in representing entities?

**Participant 4**

Difficulties remaining current

Providers anchoring on past determinations (correctly or incorrectly)

A number of disparities that would make it hard to participate (financial, housing, recent immigrants, without PCP)

Lacking information about how genetics change over time?

Issues transferring information between health systems and health record systems

How much does lifestyle matter?

Issues incidental findings and follow-up

Do we have a lens of prevention vs just acute medical conditions?

Who is going to fund the research (response to opiates* example) - historical studies

**Participant 5**

What are we going to do around minors, is this going to be shared with guardians

Flawed data going in will lead to flawed data going out

Data sharing among institutions EMRs that speak to each other

Who will pay? individual vs societal - extreme cost of care already

How are we measuring precision medicine from a quality lens (how do we know we are doing it right/well)

**Participant 6**

If we think social determinants of health are inputs, are patient values inputs?

Privacy is a concern

**Participant 7**

Oversampling some populations and undersampling others

Opportunity further racist ideology and marginalize patients - race is genetic and biologic is one argument in our country

Will precision-medicine be the same for diagnoses and populations

Expensive

How to decide what disease conditions to study and treat?

Will who we decide to study and treat have unintended consequences?

What if we are wrong (precision medicine is wrong)?

Reducing our patients to a few factors and not holistic human -- harmful to patient and doctor

How are we going to teach it?

Will all programs have access to a person who can teach this?

What about getting the right treatment for patients is not right for that human - "recommendation of admission". Can precision medicine pivot? if not inline with patients

Will non-precision medicine be available to patients or do they have to do precision-medicine pathway

**Participant 8**

How will genetic info be collected, stored, maintained? who will be accountability to this

Who will be accountable for the collected information

Provider- do we know how to interpret this information and does that change in an emergent scenario

Does how we interpret PEM change in an emergent scenario

Equity - how do we include underrepresented or excluded groups in a way that avoids surveillance

What will corporate influence in this area be?

Can different care still be equitable "separate but equal" in outcomes?

Can perception of different care still be equitable

How do we maintain humility as we learn more about genetics and epigenetics?

**Participant 9**

Should we be investing in PEM when many have no access to care

Will insurance companies access this info and use it against you

Unintended psychological consequences (person knows they will get a disease, lives in fear)

Mistrust of people/system

Parents consent on behalf of their child and they never wanted to know?

Potential lawsuits

**Participant 10**

Patient interface with own medical information and ability to interpret and apply this information

Gender issues? identity issues?

How do we ascribe value to different educational choices and the ways they impact their health care

The discoverability and legal proceeding have more information and how that impacts insurance/legality

Who owns this data?

How can we make it safe while also making it accessible

Accuracy in obtaining demographic information (is patient listing this or someone else assigning it)

**Participant 11**

How do you make sure goals of precision medicine are not being changed when going from hospital—>provider—>patient

How do we keep the interest of patient first

How do we present ideas in a way that everyone can understand?

Feedback. How do we make systems adaptable over time

How do we keep patient’s choices in precision-medicine

How do we keep the personalization in precision medicine

Social determinants: gender, financial, socially. How do we communicate this?

**Participant 12**

How do we make judgements on what is most important to prioritize

Doctors are humans too and may have judgements and values that may conflict with patients

Trust generally requires relationship built over time and how do we do that in the ED

Feedback from EM to PCP and vice versa

Genetic risk factors vs modifiable or behavioral risk factors and how to play in care

Through patient education how can we strategize to engage historically hesitant groups from participation?

Table S3: Cross-Cutting Themes

**CROSS-CUTTING THEMES**

How accurate is the data used for precision medicine?

How will payment for precision medicine be structured?

Who will have access to precision medicine?

How are we going to educate providers, patients, and the public?

Will all institutions have the capability to teach precision medicine?

Will information be transferable between health care systems?

How do we keep the interests of the patient first?

How do insurance and corporate interests fit into the model?
